# “*Another gay fear*”: community reflections on UK public health responses to the 2022 mpox outbreak

**DOI:** 10.64898/2026.02.12.26346155

**Authors:** Tom Witney, Emily Jay Nicholls, Marthe Le Prevost, Yomna Gharib, Davide Bilardi, Sarah Denford, Matthew Hamer, Parminder Sekhon, Darren Knight, Shema Tariq

**Affiliations:** Institute for Global Health, University College London, London, UK; MRC Clinical Trials Unit, University College London, London, UK; Department of Primary Care Health Sciences, University of Oxford, Oxford, UK; Health Protection Research Unit in Evaluation and Behavioural Science, University of Bristol, Bristol, UK; VERDIQual Community Advisory Board, London, UK; NAZ Project London, London, UK; George House Trust, Manchester, UK

## Abstract

**Background:** Between May and December 2022, the UK Health Security Agency reported 3,585 cases of mpox. The 2022 global outbreak was characterised by transmission predominantly within sexual networks of gay and bisexual men (GBMSM). UK public health responses included information and vaccination campaigns, supported by behaviour change among GBMSM. We describe community reflections on the UK mpox outbreak and the responses to it.

**Methods:** Between August 2023 and April 2024, we conducted five focus groups with participants (n=27) recruited through community organisations in London and Manchester. Participants were predominantly gay men from ethnically diverse backgrounds. Data were analysed thematically.

**Results:** Coming soon after COVID-19, the mpox outbreak intensified fears of returning to lockdown. Reports of GBMSM being most affected, and its framing in the media as a ‘gay disease’, coupled with warnings within GBMSM networks echoed earlier experiences of HIV. Those who had acquired mpox reported that media coverage had intensified their experiences of stigma and discrimination. Participants perceived vaccine roll out as inequitable; Furthermore, the perceived sudden cessation of public health messaging and advice left participants uncertain about ongoing risk and the need for prevention.

**Conclusions:** Participant reflections two years after the 2022 mpox outbreak demonstrate how previous pandemics shape emotional responses to new outbreaks. Key challenges included stigmatising media coverage, inequitable vaccine rollout, and sudden discontinuation of public health messaging. These findings highlight the importance of targeted, non-stigmatising and unambiguous communication from trusted sources during and after an outbreak.

**What is already known on this topic:** The media response to the 2022 UK mpox outbreak led to stigma among GBMSM and public health measures were not equitably accessible

**What this study adds:** People from communities affected by the outbreak sought timely information from trusted sources. Communication needs do not end with outbreaks

**How this study might affect research, practice or policy:** There is a need for ongoing public health work to build trusted networks who can maintain inter-outbreak communications and respond rapidly to outbreaks

## Introduction

Mpox (formerly known as monkeypox) is a zoonotic infection endemic to West and Central Africa (1), but rare outside these regions (including in the UK). Between May and December 2022, the UK Health Security Agency (UKHSA) reported 3,732 cases of mpox clade IIb (2). Unlike previous mpox outbreaks, transmission predominantly occurred within sexual networks of gay and bisexual men (GBMSM).

Initial public health responses to the 2022 mpox outbreak in the UK centred on contract tracing and isolation, later comprising targeted roll out of the modified vaccinia Ankara (MVA) vaccine. These transmission-reducing interventions were supported by public health messaging by community-based organisations, healthcare providers and official health agencies including the UKHSA. Knowledge mobilisation about mpox and vaccination within GBMSM networks and social media were crucial to managing the outbreak, with behaviour change (including reducing partner numbers, avoiding sex on premises venues, mpox testing, self-isolation, and sexual abstinence) a key driver in reducing mpox transmission (3). Modelling suggests that awareness of the outbreak and behaviour change changed the trajectory of the outbreak, with vaccination later preventing a rebound in cases (4). There has been a significant and sustained decline in mpox incidence in the UK since September 2022 with between 1 and 41 cases reported per month between 2023 and 2025, compared to the monthly high of 1,339 cases in July 2022 (2).

Qualitative investigation of experiences of the 2022 mpox outbreak are scarce. Interviews and media analyses highlight limitations in both public health and community responses. This includes poorer knowledge of and adherence to prevention measures among those outside GBMSM networks, a perceived lack of timely and accurate information from public health agencies, contradictory and inconsistent advice, and the predominance of stigmatising media narratives (5–7). These studies were conducted during or shortly after the 2022 outbreak, when the social meanings of the disease and public health measures were likely still emerging. These studies therefore capture immediate responses to the outbreak, rather than reflections on collective attitudes, social influences, and group dynamics which are developed over time (8). We aimed to explore reflections on the 2022 mpox outbreak, among GBMSM (and other people affected by mpox), to inform future health communication and strengthen sexually transmitted infection (STI) pandemic preparedness.

## Methods

This study is part of the larger VERDIQual project, which seeks to provide qualitative understandings of the 2022 mpox outbreak in endemic and non-endemic settings (9). We report findings from focus group discussions (FGDs) with participants who perceived themselves to be (or have been) at risk of acquiring mpox. We chose this approach to facilitate interaction between participants, providing insight into community knowledge, attitudes, and practices, including areas of agreement and disagreement(10).

FGDs were conducted with three community-based organisations: George House Trust in Manchester, which provides support and services to people living with HIV; NAZ, a sexual health charity focusing on sexual health in racially and sexually minoritised communities in the UK; and Positively UK, an HIV peer support charity in the UK. These organisations publicised the study within their own networks. Adverts included a link to a screening survey to which gathered demographics and contact details. The study team then contacted potential participants, sending a participant information sheet and consent form prior to the FGD.

Inclusion criteria were age ≥18 years, self-identifing as having been (or being) at risk of mpox acquisition, and ordinarily resident in the UK. We excluded people who could not speak English due to the practical difficulties of mixed language FGDs, and those who were unable to give informed consent. Participants all provided informed consent prior to taking part, and were offered a £20 gift voucher in recognition of their time and expertise.

We conducted FGDs online and in-person, lasting between 120 and 150 minutes. The topic guide focused on experiences of mpox and information-seeking, trusted sources of information, and reflections on the public health response, information campaigns and newspaper coverage (Supplementary Appendix 1). Two experienced qualitative health researchers (EJN, with either MLP or TW) facilitated each FGD. We audio-recorded and transcribed FGDs and made contemporaneous notes. We determined the number of FGDs by information power, judging when we had sufficient data to meet the study aim, balancing the exploratory research question with the specificity of the population of interest and the richness of data (11).

Data were analysed using codebook thematic analysis (12) by TW and YG, with input from EJN and MLP, supported by NVivo (version 14, Lumivero, Burlington, MA, USA). We coded FGD transcripts inductively, dual coding one transcript to develop a codebook which was then applied to all data. We discussed developing themes with our community advisory board (CAB) comprising representatives from community organisations and people with lived experience of mpox from Europe, Nigeria and Thailand. We further refined themes through these discussions and iterative engagement with data. Final consensus on themes was reached with all authors through discussion. Ethical approval was granted from the University College London research ethics committee (6698/005).

The core study team comprised infectious disease and sexual health clinicians and public health researchers who all have extensive experience of working with marginalised communities to explore sexual health and HIV needs. Members of the team have lived experience of sexual and/or racial minoritisation. The study was designed by EJN, MLP, DB and ST; FGDs were conducted by EJN, MLP and TW; data were analysed by TW and YG, with input from EJN and MLP.

## Results

We conducted five FGDs (four in person and one on-line using Microsoft Teams) with 27 participants (4–7 participants per FGD) between August 2023 and April 2024. Twenty-four participants provided demographic information (Table 1). Participants were aged between 24 and 68 years old (median age 45 years old) and predominantly identified as gay men. Four participants identified as Asian, five as Black, three as Latin, two as Mixed ethnicity, and 10 as White. Five had been diagnosed with mpox during the 2022 outbreak and 18 had accessed vaccination (including one participant who received smallpox vaccination in childhood).

**Table 1.** Participant demographics*.

|  | n | %** |
| --- | --- | --- |
| **Gender** |  |  |
| Man | 22 | 92 |
| Nonbinary | 1 | 4 |
| Woman | 1 | 4 |
| **Sexual orientation** |  |  |
| Bisexual | 1 | 4 |
| Gay | 22 | 92 |
| Heterosexual | 1 | 4 |
| **Ethnicity** |  |  |
| Asian | 4 | 17 |
| Black | 5 | 21 |
| Latin | 3 | 13 |
| Mixed | 2 | 8 |
| White | 10 | 42 |
| **Country of birth** |  |  |
| UK | 14 | 58 |
| Outside UK | 8 | 33 |
| Did not state | 2 | 8 |
| **Education** |  |  |
| No formal education | 1 | 4 |
| GCSE (or equivalent) | 2 | 8 |
| A-levels (or equivalent) | 5 | 21 |
| Degree | 15 | 63 |
| Did not state | 1 | 4 |
| **Employment** |  |  |
| Employed full time | 11 | 46 |
| Employed part time | 5 | 21 |
| Unemployed | 7 | 29 |
| Did not state | 1 | 4 |
| **Mpox diagnosis** |  |  |
| Diagnosis confirmed | 5 | 21 |
| Diagnosis suspected | 0 | 0 |
| No diagnosis | 18 | 75 |
| Did not state | 1 | 4 |
| **Vaccination status** |  |  |
| Received 1 dose | 9 | 38 |
| Received 2 doses | 8 | 33 |
| Received during childhood | 1 | 4 |
| None | 5 | 21 |
| Did not state | 1 | 4 |

\*Twenty-four participants provided demographic information. \*\*Totals may not equal 100% due to rounding

We asked participants to provide three words that they associated with mpox, collating these answers to generate a word cloud (Figure 1) which we used to sensitise us to participants’ experiences and to guide analysis. We developed three overarching themes from the data (Table 2).

**Table 2.**
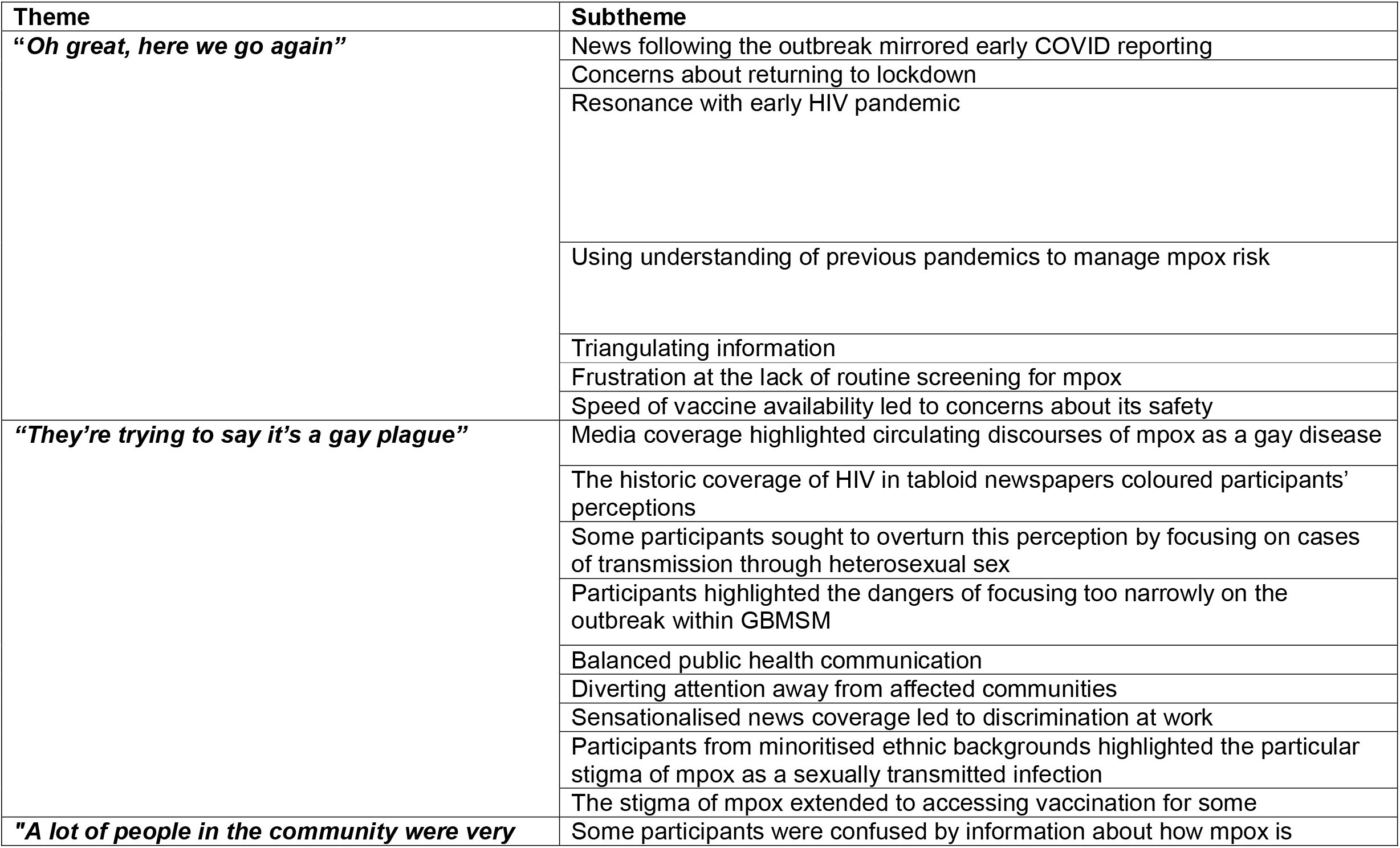
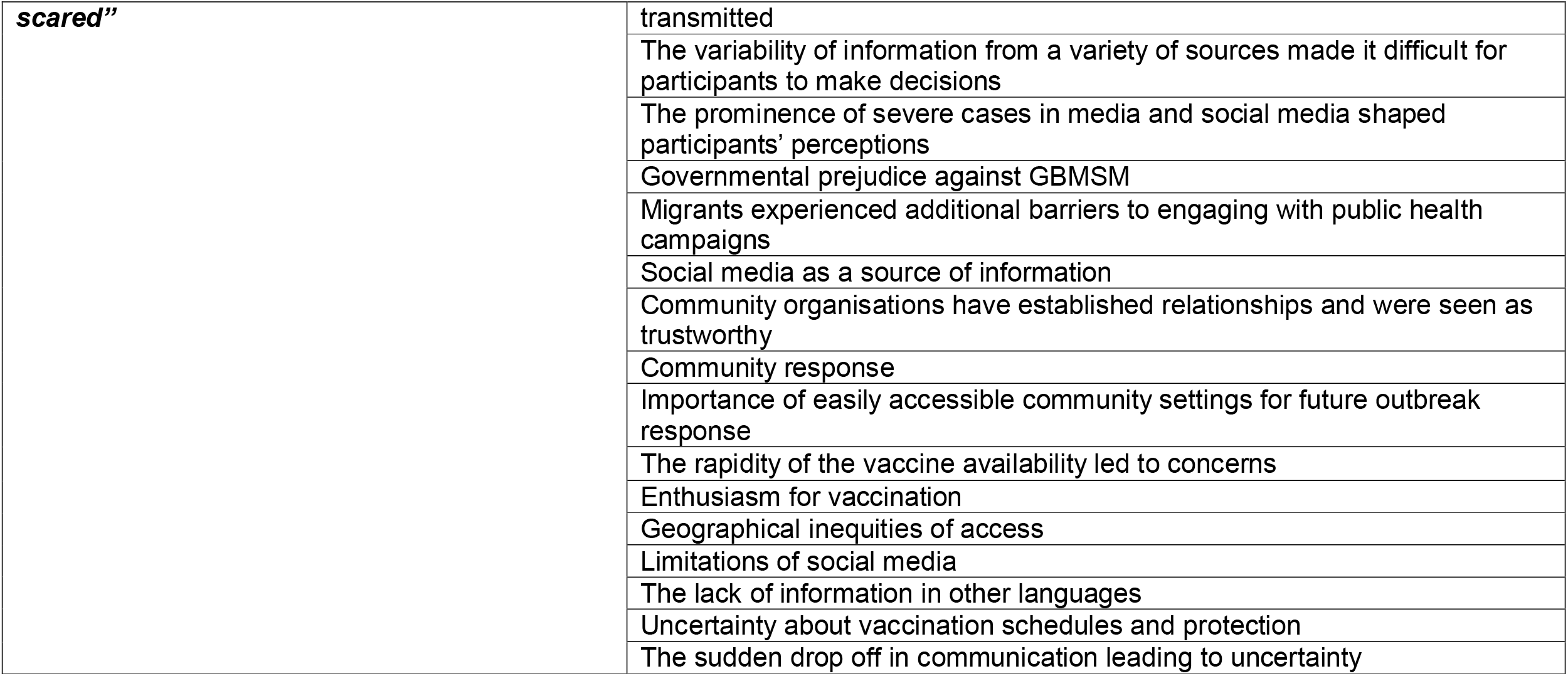
Themes and subthemes. Full details, including sample quotes are provided in Supplementary Appendix 1

**Figure 1.**
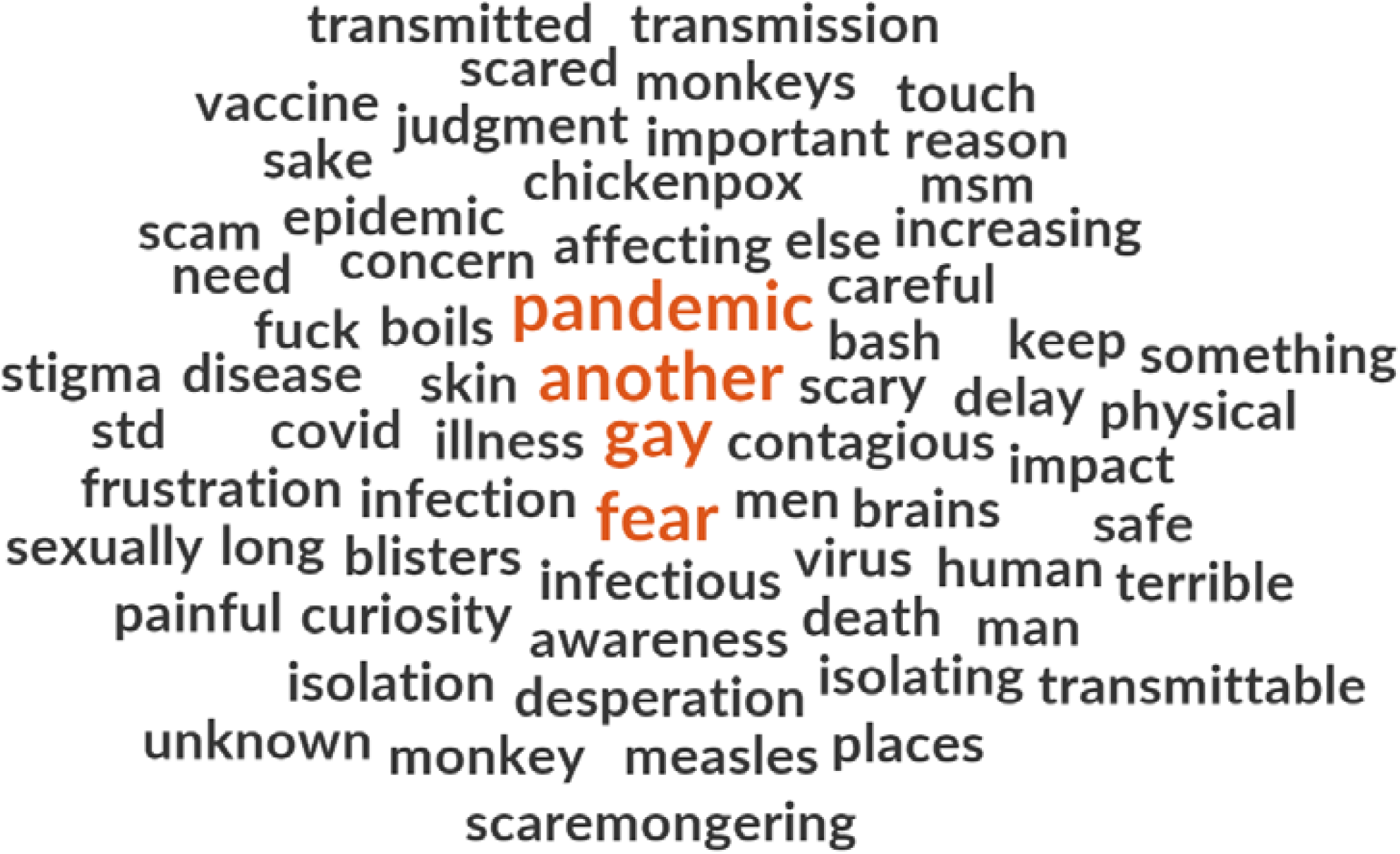
A word cloud summarising participants’ reflections on the mpox outbreak

### Oh great, here we go again

The timing of the outbreak, coming shortly after the end of COVID-19 pandemic restrictions, contributed significantly to participants’ feelings about and experiences of mpox. Their recent experiences of physical distancing and isolation during the pandemic, framed emotional responses to early news of mpox, including anxiety about potential lockdowns. Participants who had broken social distancing guidelines to meet and have sex during COVID-19 anticipated renewed feelings of guilt should social distancing return:

> *“You kind of got out of the guilt of it and you got back to normal and then suddenly you were like, you can’t do that.” Focus Group 3 participant*

Some participants first heard about mpox through friends, with rumours of an imminent surge in cases and the need to “be alert” and modify their sexual behaviour. Hearing news about mpox through friends and GBMSM networks paralleled some participants’ experiences of the early HIV pandemic, contributing to a sense that “something terrible is about to happen.”

As the outbreak progressed, some sought to understand the outbreak and their own acquisition risk through knowledge they had gained during COVID-19. They described carefully weighing-up their own susceptibility and assessing the severity of mpox and its prevalence in their local networks, when deciding whether to avoid public sex environments because “it’s just too risky to do that at the moment.” Those that did not modify their behaviour were anxious anxiety about their unmanaged risk, with one participant comparing this to his having unprotected sex before HIV pre-exposure prophylaxis (PrEP) was available. Others, understanding mpox to be less transmissible than COVID, felt reassured that they were unlikely to be at risk. Furthermore, due to the global nature of the 2022 mpox outbreak and familiarity with epidemiology principles from COVID-19, some described triangulating information about mpox and public health responses from other countries. This was often accompanied by scepticism towards UK public health responses and recommendations.

Compared with widespread advice during the COVID-19 pandemic, participants across all FGDs felt inadequately informed about mpox transmission, the evolution of the outbreak, and prevention measures. Early in the outbreak, the lack of asymptomatic screening, and limited treatment and prevention options, was a source of frustration for a population who readily engaged with STI screening and biomedical prevention technologies, such as PrEP. Once available, there was widespread enthusiasm for vaccination among participants.

### They’re trying to say it’s a gay plague

The spread of mpox through GBMSM sexual networks was a key characteristic of the 2022 outbreak. Participants perceived that mpox was consequently constructed as a gay disease in public discourse. Reviewing examples of newspaper coverage, participants highlighted how headlines sensationalised cases. A participant in Focus Group 4, who had been interviewed by a journalist described how the interviewer encouraged him to draw parallels with the early days of HIV/AIDS. Such comparisons evoked memories of historical tabloid coverage of the HIV epidemic, where blame and stigmatisation loomed large. Other participants highlighted how media coverage that foregrounded GBMSM may encourage complacency among heterosexual people, potentially placing them at risk. For those from racially minoritised communities, media coverage amplified their high levels of internalised stigma relating to their sexuality. The news adding to their sense of shame:

> *“There’s already a cultural stigma around just enjoying sex. So you throw something else like that in, you’re just adding to that stigma.” Focus Group 3 participant*

In contrast to media coverage, UK public health information campaigns from community organisations, public health bodies and local government were perceived as non-judgemental, with a focus on delivery of factual information. However, campaigns that portrayed women as well as men were seen as potentially confusing and risking not reaching those most at risk. Others critiqued the use of sexualised imagery in materials developed by some community groups as inappropriately emphasising sexual transmission, when their understanding was that mpox could be transmitted as a result of any close contact, including being in a crowded space.

Participants who had acquired mpox reported discrimination as a direct consequence of the diagnosis. For example, a participant in Focus Group 4 described being contacted by a health protection team to undertake contact tracing. He speculated that this staff member had limited experience of sexual health or GBMSM and was therefore not able to provide stigma-free, culturally competent care. The sensationalised news coverage, particularly early images of severe skin manifestations, further amplified stigma, with participants encountering fear among colleagues in their work environments when they returned following self-isolation. The stigma of mpox was also associated with its prevention; people who did not want to publicly identify as GBMSM (or as having multiple sexual partners) avoided the long queues for vaccination in clinics.

Official public health responses were hampered by perceived homophobic views among the then members of the UK government with some perceiving this as the driver behind piecemeal communication and limited vaccination availability. Stigma and discrimination manifested beyond homophobia. Those from migrant communities described how the UK’s “hostile environment” had resulted in mistrust of the newly re-named UK Health Security Agency (formerly Public Health England until January 2023), a name with connotations of border security and control, fostering suspicion and fear.

### A lot of people in the community were very scared

Participants overwhelmingly described feeling fearful during the outbreak. The profusion of information from multiple sources was sometimes confusing, leaving some feeling ill-equipped to protect themselves. Fears were heightened by the experiences of severe mpox shared on social media, with relatively limited awareness among participants of the more common and milder presentations.

Negative experiences of and attitudes to media coverage and public health agencies resulted in a lack of trusted sources of information, with many turning to social media and community-based organisations. These organisations, who had often built relationships with communities over years and had credibility, became an essential source of information and advice:

“*I know they’ve got [the] community’s best interests at heart. They’re going to give us information that is important. And that is relevant.” Focus Group 2 participant*

Others highlighted how lessons learned from HIV, such as community-led responses and support, were central to addressing fears.

The MVA vaccine was welcomed by participants as offering hope. Most were enthusiastic about vaccination, framing it as offering freedom and drawing parallels with other sexual health prevention interventions such as PrEP. Several participants accessed vaccination through outreach clinics in community-based organisations within London. A minority expressed scepticism, echoing SARS-Cov-2 concerns about the rapidity of its availability. Vaccination was difficult to access for participants outside London, causing considerable frustration at the inequitable rollout. Participants recognised there was limited vaccine supply, which led to inequity as access would often depend on whether an individual was already engaged with sexual health services, or embedded in GBMSM social media and community networks. This excluded some of the most marginalised GBMSM including those who do not read or speak English.

Having accessed vaccination, people still had unanswered questions. Participants reported being unsure of the level and durability of protection conferred by one dose of MVA. Rapidly changing policy in vaccine allocation and schedules, which differed from approaches in other countries, undermined trust in its efficacy. This uncertainty was exacerbated by what participants described as a sudden drop in public health messaging and news coverage in late 2022, a participant in Focus Group 1 reflected “*it was this big thing and then it just went*.” This left participants unsure of their ongoing risk.

## Discussion

We describe reflections among GBMSM (and other people affected by mpox) on and experiences of public health messaging, vaccination, and broader community responses to the 2022 mpox outbreak in the UK. Coming soon after the COVID-19 pandemic, the initial framing of and emotional responses to mpox were grounded in individuals’ experience of previous pandemics, including HIV. News media’s focus on the sexual transmission between GBMSM and its representation of mpox as a gay disease amplified existing homophobia, especially among those from racially minoritised communities. For those who had acquired mpox, sensationalised coverage that focused on severe manifestations intensified stigma and discrimination.

In contrast, public health communication about the outbreak was seen as balanced and non-judgemental. Mistrust of Government and official bodies undermined these messages, especially among migrant communities, and participants described seeking information from trusted sources. Community-based organisations who had fostered trusted relationships over many years with communities, were often where participants accessed support and resources.

The MVA vaccine was perceived favourably by our participants. Likening it to other sexual health interventions, vaccination offered freedom from the fear of acquiring a painful and potentially disfiguring infection and transmitting it to others. It also provided hope of avoiding the disruptions and social isolation experienced during the COVID-19 pandemic. Yet, despite the willingness to be vaccinated, some participants described significant challenges in access related to shortage of supplies, especially outside of London, and being on the margins of social and community networks in which information was being circulated.

Finally, people expressed uncertainty both during and since the outbreak, with ongoing concerns about durability of vaccine protection and their current risk of mpox acquisition. This was exacerbated by the perceived rapid cessation of communication about mpox in late 2022, and the relative absence of discussion by public health agencies since.

Many of our findings are consistent with rapid research conducted during the 2022 mpox outbreak (7). Previous work has also highlighted news media framing of mpox as a gay disease (5), the impact of mpox stigma, which often amplified pre-existing homophobia, and negative experiences of public health professionals, especially within health protection teams who lacked the experience and expertise in working with GBMSM communities (6). The centrality of fear as an emotional reaction to the outbreak and hope in vaccination, resonates experiences during the COVID pandemic (8). Participants reported high levels of engagement with prevention interventions such as behaviour modification and vaccination. Our participants’ concerns about inequitable vaccine rollout echo findings from other studies in the UK (13). Participants’ engagement with the MVA vaccine has been reported by other authors, and may reflect a wider acceptance of biomedical prevention interventions among GBMSM, such as PrEP and more recently DoxyPEP (3). Our data provides further evidence of the role of community behaviour change as a critical factor to be considered in mathematical models of mpox transmission and control (4). Crucially, through its engagement with affected communities following the end of the outbreak, our study shows how communication needs continue despite the lack of new cases, and spells out the impact of a perceived cessation of communication on community members.

Our study builds upon the scant qualitive evidence on mpox. By engaging with racially diverse GBMSM communities, we were able to include racially minoritised and migrant participants, allowing us to explore barriers to public health messaging in multiply marginalised communities. In addition, we conducted our FGDs 12–18 months after the decline in mpox cases in the UK, allowing participants to reflect on their experiences more fully. Importantly, this has highlighted how uncertainty about ongoing risk persists, even now after the public health response has largely concluded. Our sample under-represented women and nonbinary people, which limits the transferability of our findings to these communities. Additionally, recruiting through community-based organisations means that we did not reach outside of those networks.

Our findings highlight the central role of community-based organisations as trusted sources of information in outbreaks of sexually transmitted infections impacting marginalised populations. We have demonstrated the emotional impact of the 2022 mpox outbreak, which is amplified among communities who have been disproportionately impacted by previous pandemics such as HIV. Public health agencies need to recognise that communication needs do not end once incidence declines, and that ongoing public health messaging and engagement with communities is often required. We recommend establishing a national STI outbreak communication group comprising community-based organisations, public health agencies, clinicians and academics. This would facilitate timely, coordinated, consistent, and effective knowledge mobilisation to those most affected by an evolving outbreak. However, such a network will only succeed if community-based organisations are sufficiently funded in the inter-outbreak/pandemic phase, so they have the resources to continue to engage and further build trust with multiply marginalised communities.

## Conclusion

We provide further insights into experiences of infectious disease outbreaks among multiply marginalised communities. We have highlighted how previous pandemic experiences and emerging stigma shaped experiences. Where media accounts are sensationalised and potentially stigmatising, and public health agencies may not be trusted, community-based organisations become crucial to informing and supporting affected communities about evolving outbreaks. Furthermore, we show how communication needs do not end when the outbreak is judged to have ended. In an increasingly challenging financial environment, our work emphasises the importance of continued funding of such organisations to enable them to forge trusted relationships in inter-outbreak phases.

## Data Availability

All data produced in the present study are available upon reasonable request to the authors.

## Acknowledgements

We would like to thank Chris O’Hanlon from Positively UK for his support with this project.

## Author contributions

The study was designed by EJN, MLP DB, and ST. EJN, MLP and TW conducted the FGDs. data TW and YG carried out the analysis, with input from EJN and MLP. TW drafted the manuscript in consultation with ST; all authors provided critical feedback and reviewed the final manuscript.

## Supplementary appendix 1

### Qualitative investigation into mpox in endemic and non-endemic areas

#### Focus Group Discussion Topic Guide

##### Who are we and what are we trying to do?

- Thank you for agreeing to participate in this focus group discussion on mpox (also known as monkeypox).
- We are a team of experienced HIV and infectious diseases researchers from University College London.
- We are very interested to hear your opinion on health messaging during the mpox outbreak in 2022.
- The purpose of this study is to learn what you think about the messages and information from sources such as newspapers and social media as well as more official information provided by NHS clinics and governmental public health bodies during the mpox outbreak. We hope to learn things that we can use to improve communications in case of future outbreaks.

##### What will be done with this information?

- The information you give us is completely confidential. We would like to make a sound recording the focus groups so that we can capture the full discussion. The recordings will be typed up (transcribed) and then destroyed. No names will be used on the transcriptions.
- You do not have to answer any questions you’re not comfortable with and you can withdraw from the study at any time.
- We understand how important it is that this information is kept private and confidential. Participants will be asked to keep what is said within the group. However, the group setting means that we are unable to promise confidentiality.
- If you have any questions, you can always contact a study team member, or you can call the project team leaders whose names and phone numbers are on this form.
- Please check and sign the consent form to show you agree to participate in this focus group.

### Introduction

1. Welcome Introduce yourself and the notetaker and send the Sign-In Sheet with a few quick demographic questions (age, gender) around to the group while you are introducing the focus group.
2. Explanation of the process Ask the group if anyone has participated in a focus group before. Explain that focus groups are being used more and more often in health and human services research.
3. About focus groups
  - We learn from you (positive and negative)
  - Not trying to achieve consensus, we’re gathering information.
  - No virtue in long lists: we’re looking for priorities.
  - In this project, we are doing both interviews and focus group discussions. The reason for using both tools is that we can get more in-depth information from individual interview and an overall prospective on the topic of interest in focus groups.
4. Logistics
  - Focus group will last about one and a half to two hours.
  - Feel free to move around.
  - Where is the bathroom? Exit?
  - Help yourself to refreshments.
5. Ground Rules Below are the ground rules.
  - Confidentiality is paramount. Information provided in the focus group must be kept confidential. However, we cannot control what information other people share so please do not say anything you would be unhappy about being shared.
  - Be respectful and avoid having side conversations.
  - Turn off mobile phones if possible. If you need to take a call please mute yourself/leave the room.
  - People may have different experiences and opinions to you. We want to hear everyone’s perspectives and we ask you to respect any views that are different to your own. Please let us know if you think we should include any other ground rules before we start.
6. Turn on sound recorder x2.
7. Ask the group if there are any questions before we get started and address those questions.
8. Introductions
  - Go around the table and ask everyone to say who they are, why did they wanted to take part, and tell us how their day has been so far.
    - The person leading FGD the meeting should answer the icebreaker question first.
    - Listen Actively and ask follow-up questions if necessary. The goal is to unite the team and give each person the opportunity to answer.
    - Let people interact with one another.
    - Allocate enough time, and don’t rush the answers.

Suggestions:

Discussion begins, make sure to give people time to think before answering the questions and do not move too quickly. Use the prompts to make sure that all issues are addressed but move on when you feel you are starting to hear repetitive information.

### Notes

Try to avoid using words like epidemic/pandemic – want this define by the group – to us.

| Questions□ | Prompts□ |
| --- | --- |
| **1. *Sources of information (do not assume people looked for information)***□ |  |
| a. Can you please tell us three words you think of when you hear the word monkeypox/mpox?□ | • What does it mean to you?□<br>• What have you heard about it?□<br>□ |
| b. Do you remember when you first heard about mpox?□<br>□ | • Where did you first hear about it?□ |
| c. Did you have difficulty finding sources of information that you trusted?□ | • Possible Activity: post-it notes (see in relation to other activities)□<br>□ |
| d. Did you actively seek out information on mpox?□<br>□ | • Where?□ □<br>• What kind of information did you look for (e.g., how serious mpox is, how best to prevent transmission, symptoms etc)?□<br>□ |
| e. Which sources of information did you find the most useful?□ □<br>□<br>Possible Activity: Ranking Exercise□ | • Why did you find these useful?□<br>• Did you find differences in the types of information provided, based on where you found it (e.g. social media, print media etc)?□ □ |
|  | • What were the differences? ☐ |
| f. What did you think about the information available about mpox? ☐<br>☐ | • Did you had to search for information? ☐<br>• Where they easily made available? ☐ |
| **2. Attitudes towards public health messaging and policies** ☐ |  |
| a. Activity: Looking at examples provided on: ☐ ☐<br>i) newspaper article headlines/tweets/ Tik Tok videos ☐ ☐<br>ii) WHO/UK/community organisation – poster / on mpox to simulate discussion ☐ ☐ | • Before the FGD, ask people to bring their own examples and also have backups. ☐ |
| b. What do you think about the way mpox was discussed by i) and ii)? ☐ | • What were the messages advising people to do? ☐<br>• What was the tone? ☐<br>• Was the messaging for all or targeted? ☐<br>• Do you think it should have been generic or targeted? ☐ |
| c. How did they make you feel for i) and ii)? ☐ ☐ | ☐ |
| d. What do you think their effects were for i) and ii)? ☐ | ☐ |
| e. What could be done differently by i) and ii)? ☐ | ☐ |
| **3. Experiences of the 2022 mpox outbreak** ☐ |  |
| a. While we are still dealing with COVID, did you ever compare some of the mpox messaging with COVID messaging? ☐ | • Why? ☐<br>• What was the same/different? ☐ |
| b. Activity: Ranking exercise. Please rank the following infections: chlamydia, HIV, genital herpes, genital warts, gonorrhoea, mpox, public lice, syphilis, hepatitis C. ☐<br>i. Disease seriousness ☐<br>ii. Stigma ☐ ☐ | • Why? ☐<br>• What was the same/different? ☐ |
| ☐ |  |
| c. How did it feel hearing about this outbreak so soon after COVID had happened?☐ | How about HIV, was there any feeling there of going into this after HIV?☐<br>☐ |
| d. What do you know about what is happening with mpox in other countries?☐ | Africa, Asia, USA, etc.☐ |
| **4. Attitudes towards transmission-reducing behaviours (including vaccination, and in endemic settings vaccination in pregnancy)☐** |  |
| a. How important was it to you to avoid catching mpox?☐ | Did you change your behaviour when you got to know about mpox?☐ |
| b. What were your thoughts on the vaccine when you first heard about it?☐ | ☐ |
| c. How did you hear about it?☐ | ☐ |
| d. Did you or your friends get the vaccine/how did you or they access it?☐ | ☐ |
| e. Activity: Prevention☐<br>What other ways of preventing mpox were you made aware of?☐<br>☐ | Using big sheets of paper on the wall, get them to write these down.☐<br>If they do not say anything we can add these prompts reducing group sex, reducing partners, circuit break, 21-day isolation, contract tracing, healthcare seeking if symptomatic.☐ |
| f. What ways of preventing mpox would be acceptable to you?☐ | Add to sheets on walls.☐<br>If not, why?☐ |
| **5. Questions and summary section☐** |  |
| a. Does anyone have any questions about any of the discussion today or about mpox in general?☐ | 10-minute section for questions and health promotion.☐ |
| b. Highlight key points from FGD and check assumptions☐ | Ask people to add anything we missed.☐ |

### Possible extra question – but maybe better as a thread throughout

- Social and emotional impact of the mpox outbreak? - thread throughout not a separate section.

### Possible useful materials/prompts

- Ask people to bring any headlines/articles/posters they remember/stood out to them.
- Bring newspaper headlines/blow up of tweets and tick tock videos as well as public health messaging materials available in the FGD - make sure to include unhelpful/stigmatising/inflammatory examples as well as measured ones.

## Supplementary appendix 2

| Theme | Subtheme | Sample quotations |
| --- | --- | --- |
| ***“Oh great, here we go again”*** | News following the outbreak mirrored early COVID reporting | “They mentioned it on the News, someone’s got it in London. Oh, no, the same with COVID. You know, someone had it on a plane, brought it over, what’s going on, crazy.” Focus Group 1 participant |
|  | Concerns about returning to lockdown | “Well, you’d had Covid where you were just made to feel like you can’t do anything but you did, but you took the risks [to meet]. Then you kind of got out of the guilt of it and you got back to normal and then suddenly you were like, you can’t do that. And you’re like, oh great, here we go again” Focus Group 3 participant |
|  | Resonance with early HIV pandemic | “We didn’t have many cases here, it was more like in America. I was told by friends on apps, dating app, and by some gay friends, to be alert in case I would be going out with someone because that was happening. Then that just took me back to HIV in the ‘90s, feeling like something terrible is about to happen” Focus Group 3 participant |
|  | Using understanding of previous pandemics to manage mpox risk | “It’s not as contagious as COVID was or might have- You really have to have really close bodily contact with somebody to get it. And, because of that, I was just like, you know what, it’s a big ooh-aah about nothing.” Focus Group 2 participant<br>“I was self-isolating for COVID for quite a long time, so when I did come out of self-isolating and that I discovered about mpox, I thought that’s the last thing I want, so I did completely change my behaviour because I used to go to quite a lot of sex clubs and I thought it’s just too risky to do that at the moment” Focus Group 5 participant |
|  |  | “So it did kind of curtail some of my sexual activities or it did make me feel a little bit more, kind of like the way that we felt before PrEP came around, where it was like, oh, you have unprotected sex and you just go, *Shit, what happens now?*” Focus Group 3 participant |
|  | Triangulating information | “I cross-referenced a lot of information to other countries. At CDC in the States and Canada and Australia, I just cross-referenced. And, I cross-referenced information. And, yes, and read a couple of journals. But, I don't just take- I've stopped just taking everything of the NHS as the gospel truth.” Focus Group 2 participant |
|  | Frustration at the lack of routine screening for mpox | “What am I looking out for, you know, stuff like that. And, obviously with the COVID thing it was you've got a cough you get your test. There wasn't any tests.” Focus Group 1 participant |
|  | Speed of vaccine availability led to concerns about its safety | “You do feel a bit like a guinea pig. You do feel like is something going to drop off or am I going to- even though the formulation has been around for a while, but it's the fact that it's just something really new. We're just like what's going to happen in five years' time” Focus Group 3 participant |
| ***“They're trying to say it's a gay plague”*** | Media coverage highlighted circulating discourses of mpox as a gay disease | “The fear factor comes from the fact of course that the printing press can only sell their papers when they sensationalise everything” Focus Group 3 participant |
|  |  | “My friend had text me and said ‘did you hear about this?’ I was like ‘What?’, and he said ‘Oh, it's another gay thing’. I was like ‘What do you mean another gay thing?’ He was like ‘Another gay disease that's spread around now. They blame the gays, they're labelling it against us because we're sharing it sexually.’ And, it just became a bit of like a hate campaign for me” Focus Group 1 |
|  |  | “I got thrown in to it with work and they said would you go on one of the news channels and do an interview about mpox... it was that time when they said, gay and bisexual men [were] more at risk... the interviewer was really pushing towards how similar is this to HIV, how similar is this to the AIDS crisis?” |
|  |  | Focus Group 4 |
|  | The historic coverage of HIV in tabloid newspapers coloured participants' perceptions | "The business hasn't changed in 40 years. I remember my dad's Sun newspaper sitting on the kitchen table when Freddie Mercury was dying and the pictures of him. ... That there has been no change and that these kind of 'holiday hell' lines still sell and still titillate and still, you know, excite people" Focus Group 4 participant |
|  | Some participants sought to overturn this perception by focusing on cases of transmission through heterosexual sex | "That's where I don't understand why it was aimed at the gays, because apparently it came from... As I said, it was a knocking shop. It was basically a dirty great big whore party... Men and women having sex with anything, sort of thing" Focus Group 1 participant |
|  | Participants highlighted the dangers of focusing too narrowly on the outbreak within GBMSM | "If that's how things are being presented to the general public, then the general public are going to think this doesn't affect me, kind of like with HIV. Then rates will potentially go up in that part of the community as well." Focus Group 3 participant |
|  | Balanced public health communication | "I personally, didn't [feel judged]. I felt like it was ... you are currently the most affected demographic. It never felt like a judgy thing, you know, 'you should all be ashamed of yourselves', [...] It was much more of a, well this is just a reality of where you're at. This is something that is spread by skin-to-skin contact." Focus Group 1 participant |
|  | Diverting attention away from affected communities | "There were efforts to suppress originally, the idea that it was largely affecting the MSM population because people didn't want to apportion blame or didn't want the effect of that to be people blaming. And, I'm not sure- I think that backfired a bit because it meant that the most needing community weren't necessarily getting the resources, because people go, it's everybody." Focus Group 4 participant |
|  | Sensationalised news coverage led to discrimination at work | "I think the whole experience [was stigmatising]... Returning to work to an office with a sign on the door saying 'Infection'. And then having industrial cleaners come to my work, screaming 'where's the Monkeypox room' [...] Or staff treating me differently when I returned to work. Like people used to fist |
|  |  | bump you and all this sort of stuff and they wouldn't when I got back." Focus Group 4 participant |
|  | Participants from minoritised ethnic backgrounds highlighted the particular stigma of mpox as a sexually transmitted infection | "We're already made to feel bad about sex because we're not procreating, having children... You've got this complexity of sexual stigma and internalised shame from what you're given culturally. Then you have the mass media just making you feel bad." Focus Group 3 participant |
|  | The stigma of mpox extended to accessing vaccination for some | "There's people who are maybe not out or they're in a relationship and they're having extra marital stuff which their partner doesn't know about or they want to present an image, which a lot of men who have sex with other men do, about their sexual habits. This again, because it's like you will only get the vaccine if you're a total whore basically, is the message you're getting." Focus Group 3 participant |
| ***"A lot of people in the community were very scared"*** | Some participants were confused by information about how mpox is transmitted | "I had a big difficulty to understand in the beginning because I said to myself, if this is Mpox, is this only skin to skin rubbing, and it's more with gay people, then why not bisexual had the same problems, you know, what I mean, or heterosexual. Because, if something is all skin to skin rubbing, I can, obviously, hug my sister, you know. We can sit together with my girlfriend and things like that, so why can't I transmit her rather than me getting transmitted only with gay men, if it was a skin problem?" Focus Group 4 participant |
|  | The variability of information from a variety of sources made it difficult for participants to make decisions | "So it was, I don't know, I think there was a lack of consistency of how and what and where and you just got a bit like, yes. I don't think it was that easy to make a rational decision." Focus Group 1 participant |
|  | The prominence of severe cases in media and social media shaped participants' perceptions | "A friend of mine had it and he was really quite ill. And he ended up being taken into hospital. And had everything, you know, the sores and all the- You know, it's not funny but it is funny. But, looking back on it now, he's like I became the Monkeypox man. He said because I was living with HIV, and I had Monkeypox so everybody wanted to talk to me about- You |
|  |  | know, he was feeling awful. So, I kind of learned about it through his journey, which was made public because, you know, he's a gay man who is public about living with HIV" Focus Group 4 participant |
|  | Governmental prejudice against GBMSM | "There was a big part of me that was desperate for it to jump from the gay community in to... for there to be an outbreak in a primary school or a standard office environment or something like that, because there was definitely a period in the first couple of weeks that felt like that was the only way that it was going to be taken seriously by government" Focus Group 1 participant |
|  | Migrants experienced additional barriers to engaging with public health campaigns | "Who are UK Health Security? Are they like the new Public Health England or something? ... It's an awful name... A lot of public health issues affect people, and some of our service users as well, who are immigrants to the country, maybe they don't have secure work or they don't have recourse to public services... If they come to the UK and some of them have insecure immigration status, their perception is if I go to the hospital, they're going to arrest me because they know who I am." Focus Group 3 participant |
|  | Social media as a source of information | "I do follow a lot of the people who work in sexual health or LGBTQ communities on twitter, and I could see them, the frustrations they were having with a lot of the information not being kind of accurate, or misinformation being spread, and then them in turn working together to kind of have a drive to disseminate useful information and give people advise on where to get vaccines and how to get vaccines" Focus Group 5 participant |
|  | Community organisations have established relationships and were seen as trustworthy | "I know they've got their community's best interests at heart. They're going to give us information that is important. And that is relevant. And that affects us. So, for me, the community organisations were more trustworthy for me as a source of let me get what is relevant rather than government, that would sort |
|  |  | of be propaganda somehow.” Focus Group 2 participant |
|  | Community response | “I think open and honest communication as well. Not just communication, you know, between maybe a potential sexual partner, but just amongst friends. Just talking amongst your friends about this disease that will affect us or is affecting us, saying, you know, what are you doing to make sure you’re safe? Checking in on your friends, saying, have you heard about this, have you heard about, you know, the treatment, the vaccines? Just having that open and honest communication between us or amongst us will really help” Focus Group 5 participant |
|  | Importance of easily accessible community settings for future outbreak response | “You need a simpler roll-out strategy, as I said before because you should look at what is going to come next, and if you can’t learn from COVID and say, stop all these barriers that you keep putting up which cost money, and just look at effectively getting people vaccinated, which you can do through GP services, community hubs, and other services that you can set up like people have mentioned in different community places” Focus Group 5 participant |
|  | The rapidity of the vaccine availability led to concerns | “You do feel a bit like a guinea pig. You do feel like is something going to drop off or am I going to- even though the formulation has been around for a while, but it’s the fact that it’s just something really new. We’re just like what’s going to happen in five years’ time” Focus Group 3 participant |
|  | Enthusiasm for vaccination | “I think the main thing in my head was go and get inoculated as soon as you can. That was all I’d got through my head. I didn’t really read much beyond that. I was like just go and get the jab, just go and get the jab. Once I got the jab, it was just like okay, I felt a little bit better.” Focus Group 3 participant |
|  | Geographical inequities of access | “I was told [by my local sexual health service] that they didn’t even know what [mpox] was. And, they didn’t have a vaccine certainly. And, that was like, well, I’m still a gay man living here and I go to Manchester and London a lot, so why can’t I have the same access as everyone else. It just seems very |
|  |  | fragmented.” Focus Group 1 participant |
|  | Limitations of social media | “I follow a lot of the people who work in sexual health or LGBTQ communities on social media and I could see them... working together to kind of have a drive to disseminate useful information and give people advice on where to get vaccines and how to get vaccines. And then once that was done, I was impressed at how people in the community came together and said, look, we all need to go and get the vaccine now and support each other and tell each other where to get the next vaccine, or where you can find a slot to get vaccines, which is really good.” Focus Group 5 participant |
|  | The lack of information in other languages | “I have some friends. They are sex workers. They don’t know. They didn’t know what they should have known... I think that’s really something that should be considered. Let’s say for, just gay people, no, the sex workers, they are in a very, very risk, and we are not here to judge them. Most of them don’t speak English. What I’m saying, this kind of situation, I hope that mpox won’t come any more, [but] some information needs to be in other languages” Focus Group 5 participant |
|  | Uncertainty about vaccination schedules and protection | “There was a lot in the media about there’s a shortage and we might only give you one vaccination when you’re supposed to have two... There’s a lot of fear around we’re going to take the one vaccination or the one shot, will that even protect us?” Focus Group 3 participant |
|  | The sudden drop off in communication leading to uncertainty | “I get anxiety sometimes about it, because it’s like - You know, it was this big thing and then it just went. You know, there was news about it, and then it just went. And, then, it was like, okay, what next?” Focus Group 1 participant |

